# Cerebrospinal Fluid Flow within Ventricles and Subarachnoid Space Evaluated by Velocity Selective Spin Labeling MRI

**DOI:** 10.1101/2024.12.09.24318672

**Authors:** Yihan Wu, Feng Xu, Dan Zhu, Anna Li, Kexin Wang, Qin Qin, Jiadi Xu

**Author notes:** **Corresponding Author:** Jiadi Xu, Ph.D., Kennedy Krieger Institute, Johns Hopkins University School of Medicine 707 N. Broadway, Baltimore, MD, 21205. Grant support from NIH: P41EB031771, R01HL149742, R01AG080104 and R21AG074978.

## Abstract

This study aims to evaluate cerebrospinal fluid (CSF) flow dynamics within ventricles, and the subarachnoid space (SAS) using the velocity selective spin labeling (VSSL) MRI method with Fourier-transform-based velocity selective inversion preparation. The study included healthy volunteers who underwent MRI scanning with specific VSSL parameters optimized for CSF flow quantification. The VSSL sequence was calibrated against phase-contrast MRI (PC-MRI) to ensure accurate flow velocity measurements. The CSF flow patterns observed in the ventricles were consistent with those obtained using 3D amplified MRI and other advanced MRI techniques, verifying the reliability of the VSSL method. The VSSL method successfully measured CSF flow in the SAS along major arteries, including the middle cerebral artery (MCA), anterior cerebral artery (ACA), and posterior cerebral artery (PCA), with an average flow velocity of 0.339 ± 0.117 *cm*/*s*. The diffusion component was well suppressed by flow-compensated gradients, enabling comprehensive mapping of the rapid CSF flow pattern in the SAS system and ventricles. The flow pattern in the SAS system closely resembles the recently discovered perivascular subarachnoid space (PVSAS) system. CSF flow around the MCA, PCA, and ACA arteries in the SAS exhibited a weak orientation dependency. CSF flow in the ventricles was also measured, with an average flow velocity of 0.309 ± 0.116 *cm*/*s*, and the highest velocity observed along the superior-inferior direction. This study underscores the potential of VSSL MRI as a non-invasive tool for investigating CSF dynamics in both SAS and ventricles.

## Introduction

Cerebrospinal fluid (CSF) plays a pivotal role in the central nervous system, providing protection against impact, maintaining chemical homeostasis for brain function, clearing metabolic waste, and distributing nutrients. Recent insights into the glymphatic system reveal CSF’s critical part in immune surveillance (1–4) and the clearance of amyloid-beta (5–9), with implications for neurodegenerative diseases (10). However, CSF circulation dynamics are not fully deciphered, with traditional models now questioned due to advanced tracer-based imaging techniques (11–13). The interaction of CSF with the brain’s interstitial fluid via the glymphatic system, essential for metabolic waste removal and nutrient distribution, is a key focus. Dysfunctions in this system may contribute to pathological conditions like hydrocephalus and cognitive decline.(14) Recent research has revealed that the CSF circulation in the brain is more complex due to the discovery of a new meningeal layer, known as the subarachnoid lymphatic-like membrane (SLYM) in rodents (15). This membrane segregates the subarachnoid space (SAS) into outer and inner layers. Further studies using a CSF tracer (gadobutrol) and MRI have observed antegrade enhancement of the tracer along the large artery trunks (16). The enhancement appeared circumferentially around major arteries, suggesting the presence of a perivascular subarachnoid space (PVSAS). This space allows for direct, antegrade transport of the tracer along arteries and further into the adjacent cerebral cortex. Additionally, changes in periarterial molecular transport within the PVSAS were associated with reduced intracranial pressure-volume reserve capacity, particularly in patients with idiopathic normal pressure hydrocephalus. (16)

A variety of non-invasive MRI techniques have been developed to image CSF flow (17). These include phase-contrast MRI (PC-MRI) (18–20), time-of-flight (TOF) angiography (21), diffusion-weighted MRI (22–24), and intravoxel incoherent motion (IVIM) (25,26), as well as spin-labeling methods (27,28). These approaches have primarily focused on imaging CSF flow in the ventricles, where the peak flow velocity can reach up to 5 cm/s.(29,30) Detecting meaningful flow in the perivascular space (PVS) remains challenging due to the relatively slow flow velocity and limited size. The predominant approach uses a long echo time (TE) and diffusion-weighted MRI sequence to calculate the pseudo-diffusion coefficient of the PVS and SAS around arteries (22,23). However, this method is difficult to differentiate between CSF diffusion and pulsatile CSF motion. Despite significant efforts dedicated to studying CSF flow in the ventricles and PVS, it remains unclear how CSF is transported within the large cavity of the SAS. The discovery and study of flow dynamics in the PVSAS still rely heavily on the highly invasive intrathecal injection of tracers (16). In theory, PC-MRI-based flow mapping can be used to visualize CSF flow in the SAS. However, most phase-contrast and spin-labeling methods are limited to single-slice imaging. When aiming to selectively and noninvasively visualize the whole SAS system, the 3D imaging method is still preferred.

Velocity selective spin labeling (VSSL) has utilized the Fourier-transform-based velocity selective pulse train for labeling slow flow while also suppressing the imbalanced diffusion attenuation by using a flow-compensated control (31,32). Fourier-transform-based VSSL pulse trains were designed by concatenating a series of small flip-angle RF pulses, interleaved with paired refocusing pulses and velocity-encoding gradients. These advanced VSSL pulse trains have been effectively employed in different MRI sequences for magnetic resonance angiography (MRA) (33–39), quantitative mapping of blood flow (31,40–48) and blood volume (32,49–51), as well as venous oxygenation (52).

The objective of this work is to propose a non-invasive VSSL MRI technique to selectively visualize and evaluate CSF flow dynamics within the SAS and ventricular systems,. We will fine-tune the VSSL module specifically for CSF flow quantification. Calibration of the flow velocity will be conducted via PC-MRI, and a detailed examination of the flow direction in both the PVSAS and ventricles is planned. This quantitative method provides a novel tool for understanding CSF dynamics in brain development as a promising diagnostic marker candidate for diseases.

## Methods

### Participants

A total of 14 healthy volunteers (age: 39±17 years; 6 females, 8 males) participated, and the specific number of participants for each study is detailed below. Scanning procedures were conducted on a Philips MR Ingenia Elition 3.0T scanner (Philips Healthcare, Best, The Netherlands), utilizing a quadrature body transmit coil and a 32-channel receive head coil. Ethical approval was obtained from the Johns Hopkins Medicine Institutional Review Board (IRB), and all participants have provided their informed consent.

### VSSL sequence

VSSL pulse train was used in this study, consisting of 9 excitation pulses (20° hard pulses), interleaved with pairs of refocusing pulses (180° composite pulses) and triangular gradient lobes of alternating polarity with 8 velocity-encoding steps (Figure 1A) (31). The control modules used a velocity-compensated gradient configuration for a more balanced diffusion-weighting effect ( *b*_*label*_ = 3.57 *s* ∕ *mm*^2^, *b*_*control*_ = 1.36 *s* ∕ *mm*^2^)(53). Images were then acquired following a post-labeling delay (PLD) of 10 ms to minimize the possible exchange of labeled blood water with CSF.

**Figure 1.**
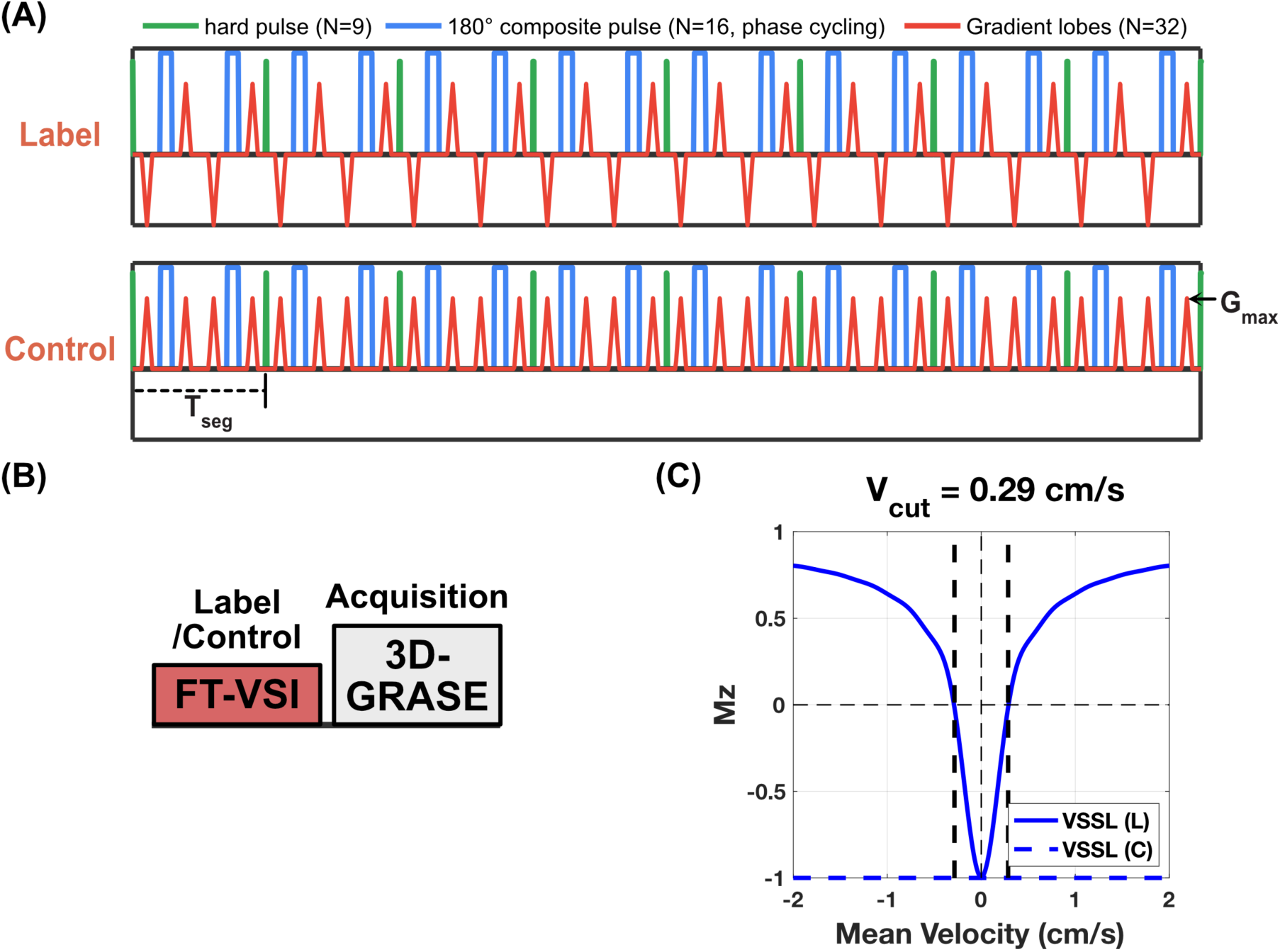
Illustration of the Fourier-transform-based velocity selective inversion pulse train, VSSL sequence, and the VSSL magnetization as a function of mean velocity. (A) Diagram illustrating the Fourier-transform-based velocity selective inversion pulse train, where velocity-selective labeling is employed to mark the flow within CSF. In each velocity-encoding step, a 20° excitation pulse is followed by a pair of refocusing pulses. Gradients with alternating polarity surround the refocusing pulses for the velocity-sensitive waveform (label), and uni-polar gradients are used for the velocity-compensated waveform (control) in the velocity-encoding steps. The segment time (T_seg_) and maximum gradient (G_max_) for the flow encoding are labeled in the sequence. (B) Diagram illustrating the VSSL sequence. After the velocity selective inversion pulse train, a post-labeling delay of 10 ms precedes the readout. A 3D-GRASE readout with a long TE is applied to image CSF while attenuating signals from parenchyma and blood. (C) Simulated Mz-velocity responses for the VSSL pulse train. Velocity-sensitive and velocity-compensated profiles are represented by solid and dashed blue lines, respectively. The cut-off velocity (V_cut_) is delineated at the first intersection where ΔM = 1, highlighted by vertical black dashed lines at a velocity of 0.29 cm/s. Parameters used in the simulation, such as number of segments (8), T_seg_ (30 ms), and G_max_ (40 mT/m), are consistent with those employed in our study.

A 3D-gradient and spin echo (3D-GRASE) readout with linear order was used for image acquisition. The MRI signals from the parenchyma and blood are attenuated by a long TE, taking advantage of their much shorter T_2_ relaxation times (<100 ms) than that of CSF (>1000 ms) (54). The imaging parameters were as follows: field of view = 220×165×160 mm^3^; slice number 80; acquisition matrix 112×82; reconstruction matrix size 224×224; acquisition resolution = 2×2×2 mm^3^; turbo spin echo (TSE) factor = 41 with linear ordering; echo-planar imaging (EPI) factor = 41; SENSE factor (S direction) = 2; echo spacing = 40 ms; TR/effective TE = 10 s/816 ms. An alternative 3D-GRASE readout changing to centric order and effective TE = 39 ms was also evaluated for comparison. We used two-sample t-tests to test whether there is a significant difference between the signal acquired from TE = 816 and 39 ms.

The control and label experiments were repeated six times for averaging, with a total scan duration of 5 minutes for each of the three orthogonal velocity-encoding directions (S-I: superior-inferior; A-P: anterior-posterior; L-R: left-right). M0 image was acquired without VSSL labeling and a TR of 20 s to ensure full recovery of CSF magnetization, other parameters are identical to the VSSL scan.

### Simulation

Simulations were performed using MATLAB 2023b (MathWorks, Natick, MA, USA). VSSL module was evaluated through Bloch simulations, including 240-ms duration with eight 30-ms segments, 16 refocusing pulses (1.74 ms) with MLEV-16 phase cycling, 0.6 ms gradient lobe duration with 0.3 ms ramp time, and maximal gradient strength of 40 mT/m, as illustrated in Figure 1A. The cutoff velocity (V_cut_) as defined in the VSASL guideline paper (55) was set at 0.29 cm/s. Mz values from label and control were simulated across velocities ranging from −2.0 cm/s to 2.0 cm/s at 0.01 cm/s intervals. In the simulation, a laminar flow model was assumed for simplification. T_1_ and T_2_ effects were not included in this simulation.

### VSSL Optimization for CSF flow

Given that the T_1_ and T_2_ values of CSF significantly differ from those of brain tissue and blood, the parameters used in MRA and CBF studies are unsuitable for CSF flow mapping. Utilizing a longer T_seg_ reduces the cutoff velocity, which is beneficial for detecting slow CSF flow. However, a longer T_seg_ also prolongs the total duration of the VSSL module, potentially diminishing the overall signal due to T_2_ decay. In regions with minimal flow motion, T_2_ relaxation becomes the dominant factor, while the VSSL will increase in regions with a wide range of flow rates. Therefore, it is crucial to optimize T_seg_ to maximize the CSF signal, while G_max_ was set to the maximum value of 40 mT/m for the current scanner. Three volunteers (30∼56 years old; 2 females, 1 male) have undergone the study of optimizing the VSSL module for CSF imaging. To optimize the VSSL pulse train targeting the CSF flow around arteries, and flow in PVS and ventricles, we varied the duration of each velocity-encoding segment (T_seg_). We fixed gradient strength (G_max_) of 40 mT/m, ramp time of 0.3 ms, and a lobe of 0.6 ms, while adjusting T_seg_ as 10, 20, 30, 40, and 50 ms (V_cut_ = 0.86, 0.43, 0.29, 0.21, and 0.17 cm/s, VSSL module duration = 80, 160, 240, 320, 400 ms, respectively).

### PVSAS and ventricle assignment

To identify the signal from PVS and ventricles or the CSF flow around the arteries in the SAS, images acquired with VSSL preparation were overlapped with velocity-selective MRA and T_1_ weighted (T_1w_) anatomical images from MPRAGE. MRA was obtained with a Fourier-transform-based velocity selective saturation module followed by a gradient-echo (GRE) readout (56). The VSSL pulse train applied 9 excitation pulses (10° hard pulses), T_seg_ = 10 ms, G_max_ = 30 mT/m, gradient lobes of 0.6 ms with ramp time of 0.2 ms. MRA scan used identical geometry as those VSSL scans including the field of view, resolution, and orientation. For the GRE readout, resolution = 1×1×1 mm^3^, TR/TE = 9.1/1.8 ms, flip angle = 8°, TFE factor = 90 with a centric ordering, and compressed sensing factor = 8 was used. The MPRAGE sequence parameters were as follows: field of view = 220×200×200 mm^3^; resolution = 1×1×1 mm^3^; TR/TE = 7.9/3.7 ms; TFE factor = 128 with a linear ordering; flip angle = 8°, SENSE factor (P direction) = 2. The total scan duration for MRA and MPRAGE were 1 min and 3.5 min, respectively.

### Calibration of the CSF flow measured by VSSL

Eleven volunteers (22∼79 years old; 4 females, 7 males) were involved in the calibration study. As demonstrated in Figure 1C, the VSSL signal intensity can be approximated as a linear function of the flow velocity when the flow is slow. Consequently, we calibrated the VSSL-derived velocity maps to actual flow velocity using PC-MRI. The sagittal plane that covers the CA, 4V, and SC was localized using a sagittal MPRAGE scan. Retrospective cardiac-gated 2D phase contrast MRI scans were performed using a 2D single-slice GRE sequence with bipolar velocity-encoding gradients. Each experimental set comprised phase contrast data acquisition at flip angles of 20°, with the remaining MR parameters being a field of view of 220×160 mm^2^, an in-plane resolution of 0.5×0.5 mm^2^, a slice thickness of 4 mm, TR/TE of 30/20 ms. The velocity encoding (VENC) was set at 10 cm/s across 16 cardiac phases with the flow direction of S-I and at 3 cm/s with the flow direction of A-P and L-R. Physiological waveform monitoring was conducted using a Philips Peripheral Pulse Unit. To correct for background phase offset, all phase images from a cardiac cycle were first averaged to produce a mean phase image. The mean phase image was then processed using a median filter with a 50×50 kernel size. Finally, the filtered mean phase image was subtracted from the phase image of each cardiac phase. (57)

### Data analysis

All analyses were processed using custom-written MATLAB scripts. The VSSL images were obtained by subtracting the control and labeled images in a pairwise fashion using a complex subtraction, and subsequently normalized by the M0 signal:

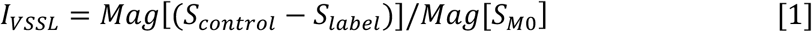

*S*_*control*_, *S*_*label*_, and *S*_*M*0_ represent the control, label, and M0 images in complex value, respectively. The *Mag*[⋅] operator takes the magnitude of a complex value. *l*_*S*–*l*_, *l*_*A*–*P*_, and *l*_*L*–*R*_ represent the normalized VSSL signal acquired with S-I, A-P, and L-R velocity-encoding direction, respectively. ROIs representing CSF within interpeduncular cistern (IPC), prepontine cistern (PPC), and spinal canal (SC) regions were manually delineated on the PC-MRI images to determine flow rates. The VSSL-derived values, extracted using the same ROIs, were correlated with flow velocities obtained from PC-MRI. As VSSL measured the average CSF velocity, the average absolute velocity measured by PC-MRI in one cardiac cycle was used for the calibration.

To access the CSF movement around the arteries in the SAS, we depicted the surroundings of the cerebral arteries shown on MRA. We chose three arteries for the SAS analysis, including M1 and M2 segments of the middle cerebral artery (MCA), A3 segment of the anterior cerebral artery (ACA), P1 and P2 segments of the posterior cerebral artery (PCA). The ventricular ROIs were manually delineated on individual participants’ M0 images, including the third ventricle (3V), fourth ventricle (4V), and frontal horn of the lateral ventricle (FLV), cerebral aqueduct (CA), and foramen of Monro (FMo). IPC and SC were also included in our analysis.

To determine whether there are statistically significant differences among the CSF velocities of three orthogonal directions (S-I, A-P, and L-R), we performed a one-way analysis of variance (ANOVA). Following the ANOVA, where a significant F-statistic indicated differences among group means, post-hoc pairwise comparisons were conducted using Tukey’s Honest Significant Difference (HSD) test. We considered a *p*-value of less than 0.05 to be statistically significant.

## Results

### Simulation

Figure 1C illustrates the simulated Mz-velocity responses for the VSSL pulse train with laminar flow integration. At a flow velocity of 0 cm/s (stationary spins), the magnetization is fully inverted by the VSSL pulse train, whereas at flow velocities exceeding 0.29 cm/s, magnetization is partially attenuated (0 to 0.8 of Mz). Below 0.8 cm/s, the Mz displays a nearly linear relationship with the flow velocity. However, CSF exhibits complex flow patterns in the brain, characterized by a mixture of various orientations and flow velocities, making it challenging to simulate. The simulated Mz-velocity responses for the VSSL pulse train with varying segment times are shown in Supplementary Figure S1 for comparison.

### Optimization of T_seg_

Figure 2 illustrates the T_seg_ optimization of the VSSL pulse train for the CSF flow images. Figure 2A exhibits the VSSL images from a typical subject with varied T_seg_ ranging from 10 ms to 50 ms. The image quality shows an obvious improvement when T_seg_ is increased from 10 ms to 30 ms, as evidenced by the highly reduced background with longer T_seg_. The background signal may partially originate from the PVS, but this requires further validation. Since the PVS signal is much weaker than the CSF signal in the ventricles and SAS, it is challenging to study and will be addressed in future research. The VSSL signal decays slowly as a function of T_seg_ in most regions. Figure 2B shows the normalized VSSL signal around the MCA, ACA, PCA, and ventricular regions including 4V, CA, and FLV. The signal around MCA and ACA remains relatively stable across T_seg_. The signal around PCA, 4V, and CA decreases with T_seg_. Notably, the signal in the FLV, considered as “free-water” with minimal motion (58), decreases from T_seg_ = 10 ms to 30 ms but stabilizes from T_seg_ = 30 ms to 50 ms. The signal in FLV can be used to assess the contribution of the diffusion component. With T_/seg_ = 30 ms, the normalized VSSL signal in FLV is only 0.006, which is far less than (<25%) the normalized VSSL signal around MCA (0.050), ACA (0.028), and PCA (0.070). Thus, T_seg_ = 30 ms was selected for further analysis due to better image quality, low diffusion-weighted effect, and relatively high VSSL signal.

**Figure 2.**
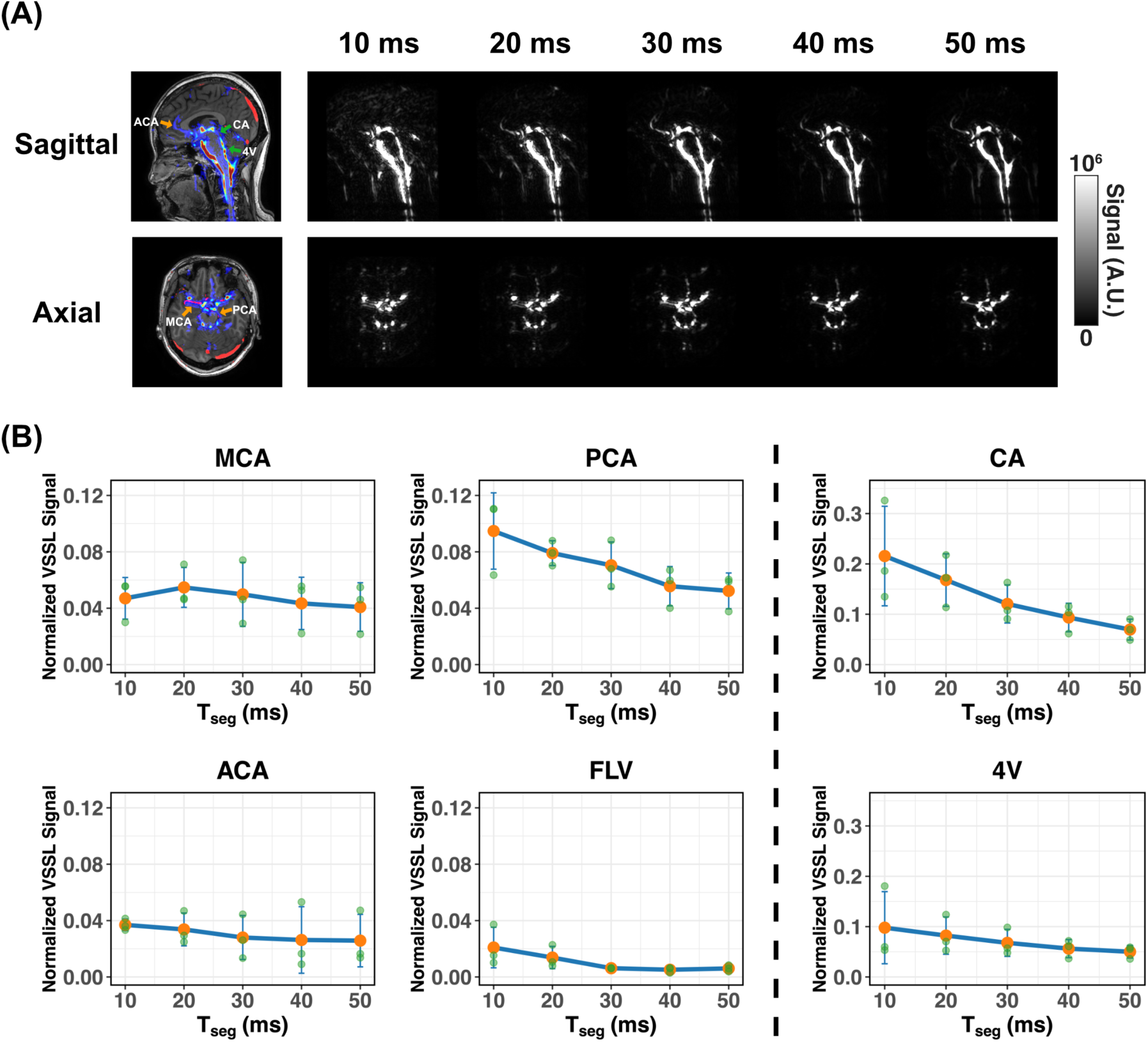
T_seg_ Optimization. (A) Example VSSL images from one subject acquired with different T_seg_ in sagittal and axial views. The left images, overlapped with MRA and MPRAGE images, show the ROIs for the VSSL signal presented below. (B) Normalized VSSL signal as a function of T_seg_ around MCA, ACA, and PCA, as well as at the 4V, CA, and FLV (N = 3, error bars represent standard derivation). The normalized VSSL signal around MCA and ACA are relatively stable across different T_seg_, while the signal decreased significantly from T_seg_ = 10 to 30 ms in the CA, 4V, FLV, and around PCA. The arteries in SAS are indicated with yellow arrows, and other ROIs are labeled with green arrows.

### The VSSL signal as a function of TE

Figure 3A presents representative normalized VSSL images acquired at TE of 816 ms and 39 ms for a subject. The velocity-encoding gradient was oriented along the S-I direction. The normalized VSSL signals around the arteries in the SAS and in the ventricles were plotted in Figure 3B and summarized in Table 1. The signals are substantially higher at TE = 39 ms compared to TE = 816 ms, with their signal ratios ranging from 1.14 (CA) to 2.09 (3V) (Table 1). For the signals around the arteries in SAS, the signal ratios ranged from 1.23 (A3) to 1.91 (P1). Although the normalized VSSL signals at TE = 39 ms are overall higher than those at TE = 816 ms with less T_2_-decay effect, significant differences were only found in P1 and P2 segments of the PCA, IPC, and SC, which is expected for CSF with a long T_2_ value. When using the VSSL module, a long Tseg (30 ms) was applied, resulting in an extended VSSL preparation time (270 ms), which effectively suppressed both parenchyma and blood signals. The M0 images acquired with a short TE were able to detect weak tissue signal as demonstrated in the Supplementary Figure S2, likely due to the exceptionally long acquisition time, which led to blurring and reduced SNR for the short T_2_ components. Despite the enhanced signal at TE = 39 ms, the image quality at TE = 816 ms was superior. The centric ordering used for TE = 39 ms may result in contamination from tissue in the M0 images (Fig. S2).(59) Therefore, we utilized TE = 816 ms for the acquisition.

**Figure 3.**
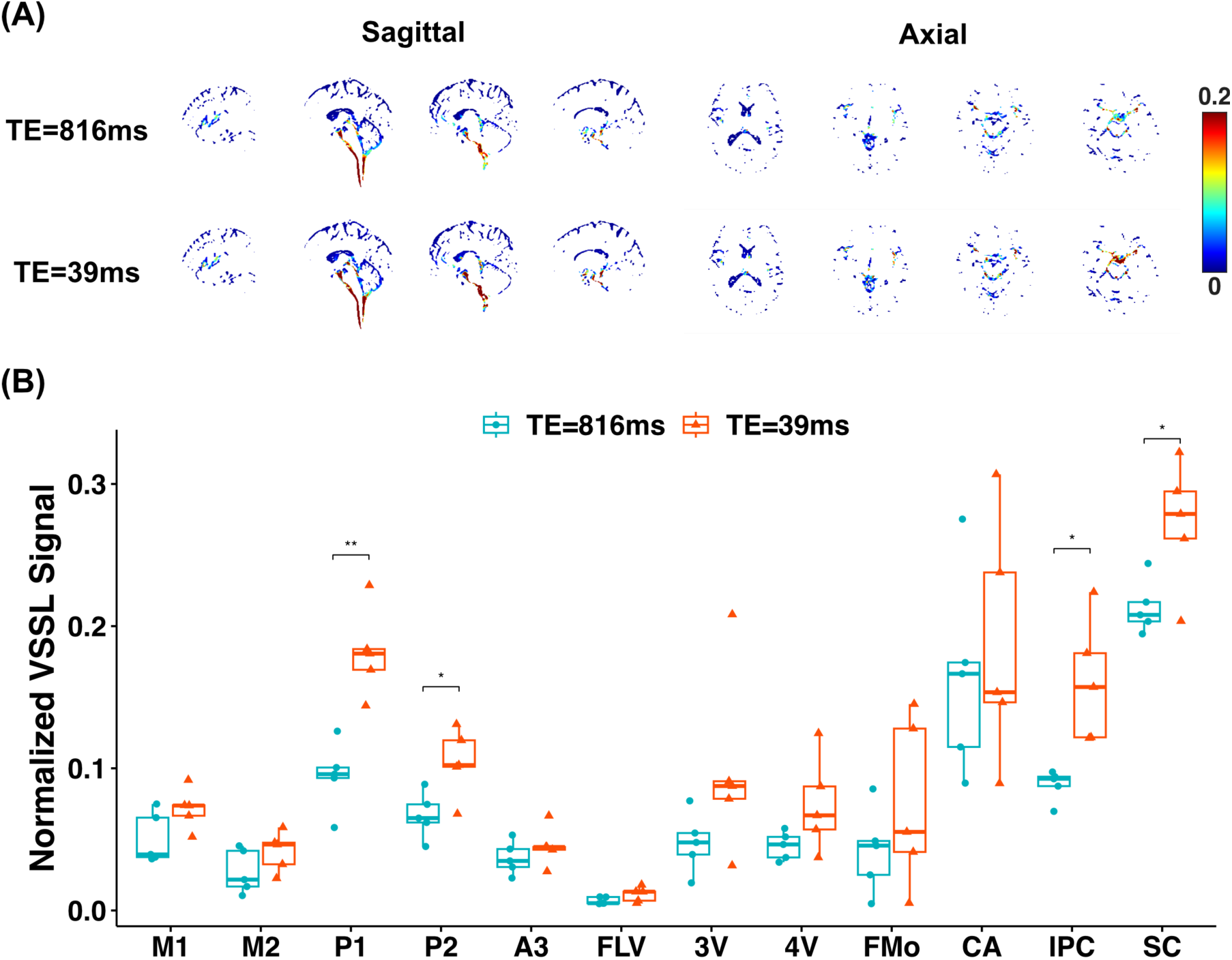
The results for the normalized VSSL signal with different TEs. (A) The typical normalized VSSL images acquired at TE = 816 and 39 ms with S-I velocity-encoding gradient directions for a representative healthy subject. (B) The boxplot of normalized VSSL signals in each ROI with different TE. A two-sample t-test was performed for comparison (* *p* < 0.05, ** *p* < 0.01).

**Table 1.** The normalized VSSL signal, signal ratio, and *p*-value with signal acquired at TE = 816 and 39 ms for each ROI. A two-sample t-test was used to measure whether there is a significant difference between the normalized signals acquired at TE = 816 and 39 ms (N = 3).

| ROI | M1 | M2 | P1 | P2 | A3 | FLV | 3V | 4V | FMo | CA | IPC | SC |
| --- | --- | --- | --- | --- | --- | --- | --- | --- | --- | --- | --- | --- |
| TE = 816 ms | 0.051 | 0.027 | 0.095 | 0.067 | 0.037 | 0.007 | 0.048 | 0.045 | 0.042 | 0.164 | 0.088 | 0.213 |
| TE = 39 ms | 0.072 | 0.042 | 0.181 | 0.104 | 0.045 | 0.011 | 0.099 | 0.075 | 0.075 | 0.187 | 0.161 | 0.272 |
| Signal Ratio | 1.413 | 1.521 | 1.912 | 1.557 | 1.225 | 1.663 | 2.086 | 1.642 | 1.786 | 1.138 | 1.824 | 1.276 |
| *p*-value | 0.080 | 0.168 | 0.001 | 0.023 | 0.337 | 0.134 | 0.155 | 0.122 | 0.311 | 0.662 | 0.018 | 0.038 |

### CSF flow patterns in SAS and ventricles

The CSF flow distribution revealed by the VSSL method is presented in Figure 4. The M0 image is shown in Figure 4A. The images, representing the sum signal of three orthogonal velocity-encoding directions 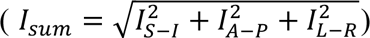, are depicted in Figures 4B-D. The average of VSSL images acquired from three velocity-encoding directions (*S*_*average*_ = ∑_*S*–*l*,*A*–*P*,*L*–*R*_(*S*_*control*_ − *S*_*label*_) /3) is overlaid with the MRA map and the MPRAGE image to identify the arteries in SAS and ventricles (Figures 4E-G). CSF flow around the arteries of M1 and M2 segments of MCA, A3 segment of ACA, and P1 and P2 segments of PCA was detected.

**Figure 4.**
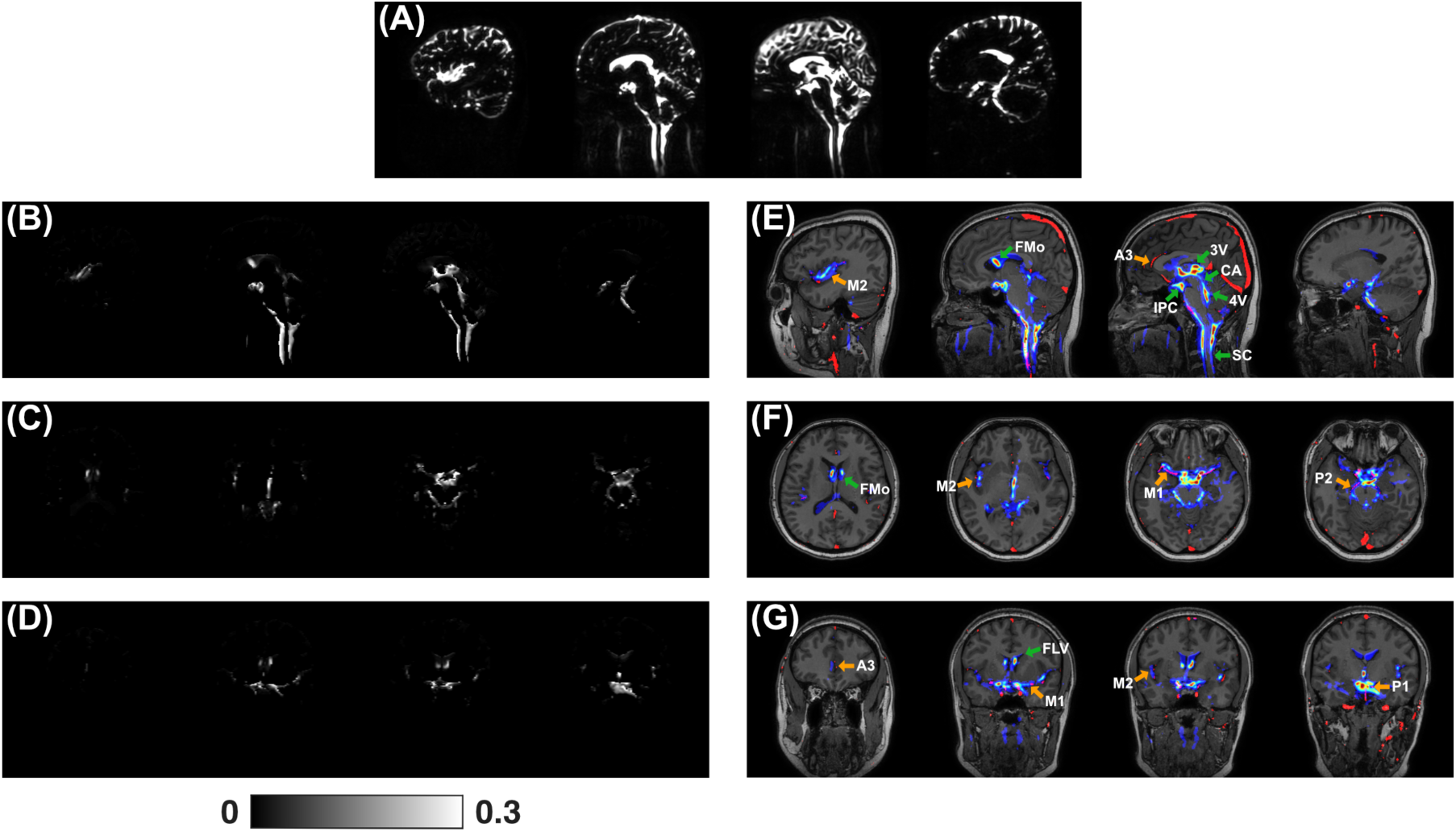
The M0 map (A) and normalized VSSL images (*l*_*sum*_) in sagittal (B), axial (C), and coronal (D) views. VSSL images were acquired with T_seg_ = 30 ms and G_max_ = 40 mT/m. VSSL image (*S*_*average*_) was overlaid with the MRA map and MPRAGE image in sagittal (E), axial (F), and coronal (G) views. The MRA map is shown in red, and the unnormalized VSSL signal is displayed using a jet colormap. The arteries in SAS are indicated with yellow arrows, and other ROIs are labeled with green arrows.

To better visualize the velocity distribution both around the arteries in SAS and in ventricles, maximum-intensity projections (MIP) are shown in Figure 5 with three views. These MIP images were produced directly from the VSSL image *S*_*average*_ without any manual enhancement. The VSSL signal in the SAS, especially in the cortical sulci region as demonstrate in the Supplementary Figure S2, is not visible, indicating effective suppression of the diffusion component in the VSSL images (Figure S3). The CSF flow around the arteries of the PCA and MCA is much higher than in other arteries. Rapid CSF flow is mainly observed in the SC and IPC, as well as in ventricles, including the 3V, and 4V. The CSF flow in the CA and FMo is also measured, further validating the VSSL method.

**Figure 5.**
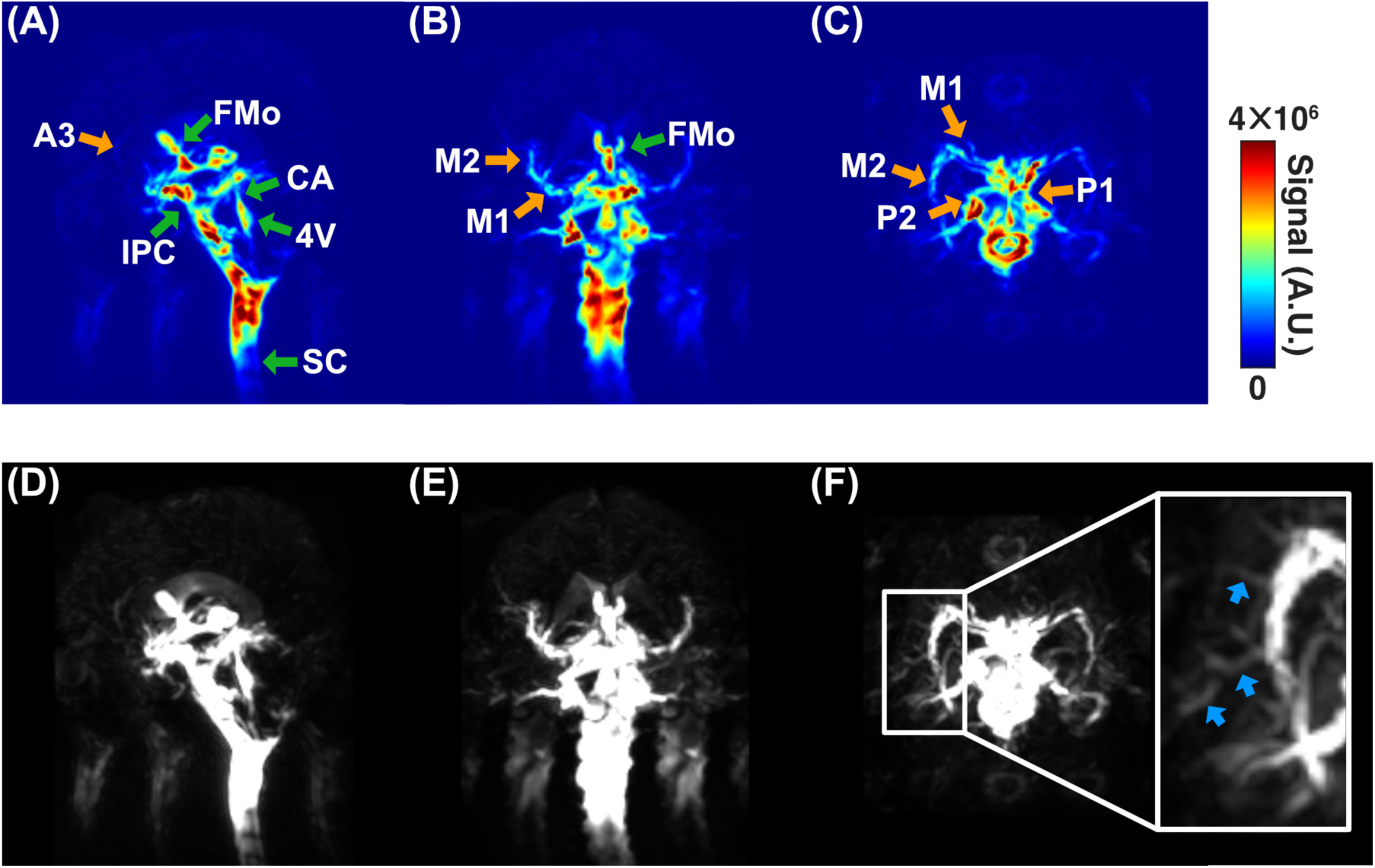
Maximum-Intensity Projections (MIP) of the VSSL image (*S*_*average*_) in sagittal (A, D), coronal (B, E), and axial (C, F) views. The top row images (A-C) use a jet colormap to show the VSSL signal in the SAS and ventricles. The bottom row images (D-F) employ a zoomed scale to highlight the small arteries, as shown by blue arrows in the inset figure. The arteries in SAS are indicated with yellow arrows, and other ROIs are labeled with green arrows.

The VSSL method effectively detects CSF flow around the major intracranial arteries (MCA, ACA, and PCA). However, detecting the flow around the basilar artery (BA) is challenging due to the extremely high CSF flow in the PPC, which obscures the CSF flow signal around the BA. The MIP images demonstrate that rapid CSF flow is confined around the MCA, ACA, and PCA, as the signals in the adjacent SAS are relatively low.

### Flow velocity calibration with PC-MRI

The CSF movement in the IPC, PPC, and SC were successfully detected using 2D high-resolution PC-MRI in all subjects. Figure 6A illustrates the imaging position of PC-MRI and the detected flow map with VENC of 10 cm/s along the S-I direction from one exemplary subject. The normalized VSSL signal acquired with S-I velocity-encoding direction is plotted in Figure 6B from the same subject for comparison. Figure 6C displays the PC velocity and PC absolute velocity waveform of the ROI shown in Figure 6A for illustration. The pulsatile flow pattern can be clearly visualized. The velocity waveforms were also extracted from all detected ventricles, which showed consistent pulsatile flow patterns across a cardiac cycle. Figure 6D shows the correlation between the normalized VSSL signal and average PC velocity of the whole cardiac cycle values measured by PC-MRI in all three directions, which shows a clear linear correlation. We applied a linear regression model with no intercept term to evaluate the correlation of normalized VSSL signal intensity and PC velocity, and then obtained the velocity maps.

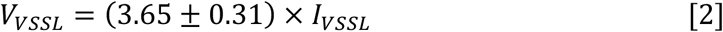

*V*_*S*–*l*_, *V*_*A*–*P*_, and *V*_*L*–*R*_ represent the obtained velocity along S-I, A-P, and L-R direction, respectively. The absolute velocity is calculated by 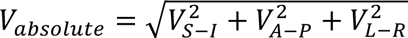.

**Figure 6.**
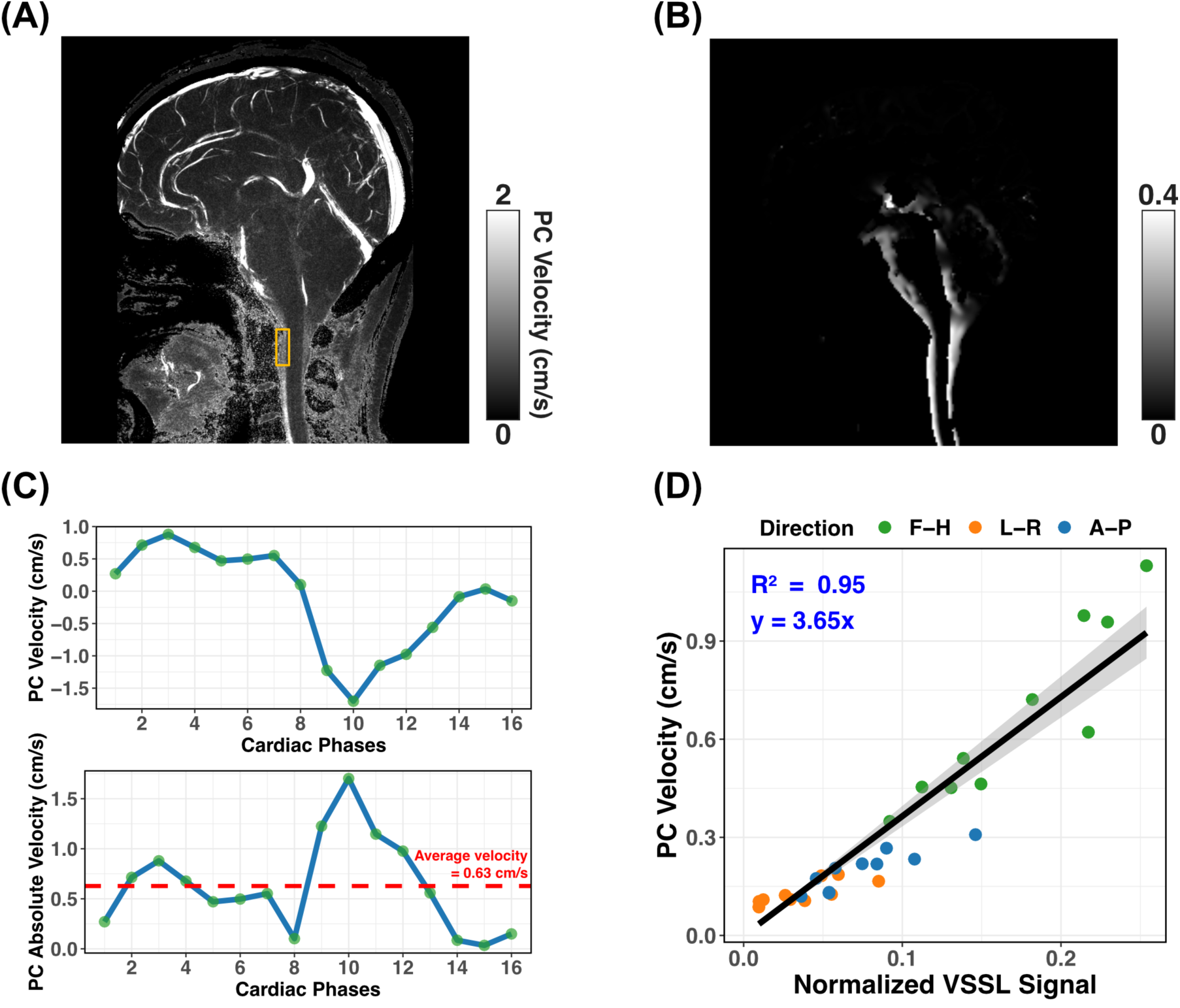
Calibration of VSSL with PC-MRI. (A) Velocity map obtained with PC-MRI for a representative healthy subject, showing pulsatile CSF flow throughout the SC, IPC, and blood flow in vessels. (B) Typical normalized VSSL images acquired with the S-I velocity-encoding direction on the same subject. (C) Representative PC flow curves with the S-I velocity-encoding direction for one cardiac cycle measured with pulse gating from the ROI shown in Figure A (yellow box). The procedure for calculating the average flow rate is illustrated in the image below. This involves taking the absolute values of the dynamic flow rates over cardiac cycle and then computing their average to obtain the final result. (D) Linear regression model between normalized VSSL signal and average velocity of the whole cardiac cycle in all three directions for the manually selected ROIs in the IPC, PPC, and SC. *R*^2^ = 0.95.

### The CSF flow along different directions

The typical velocity maps derived from Equation [2] with velocity-encoding gradients along S-I, L-R, and A-P directions and the corresponding boxplots are shown in Figure 7. The VSSL-derived velocities from different ROIs are listed in Table 2. CSF flow around M1, P1, and P2 exhibit high flow velocities of 0.461 ± 0.143 *cm*/*s*. The CSF velocities around M2 and A3 are much slower (0.155 ± 0.062 *cm*/*s*), but the flow velocity is still about 3 times higher than that of the FLV (0.041 ± 0.013 *cm*/*s*). Significant orientation dependence was observed around M2 and A3 (*p* < 0.05), with the highest velocity along the vessel walls. While CSF flow around other arteries also show a consistent trend of the highest velocity along the vessel walls, it does not reach statistical significance. The average CSF flow velocity in ventricles is 0.309 ± 0.116 *cm*/*s*. A significantly higher flow velocities along the S-I direction are found in most ventricular regions compared to those of A-P and L-R directions. The CSF flow velocities are 0.525 ± 0.105 *cm*/*s* in IPC and 0.819 ± 0.157 *cm*/*s* in SC.

**Figure 7.**
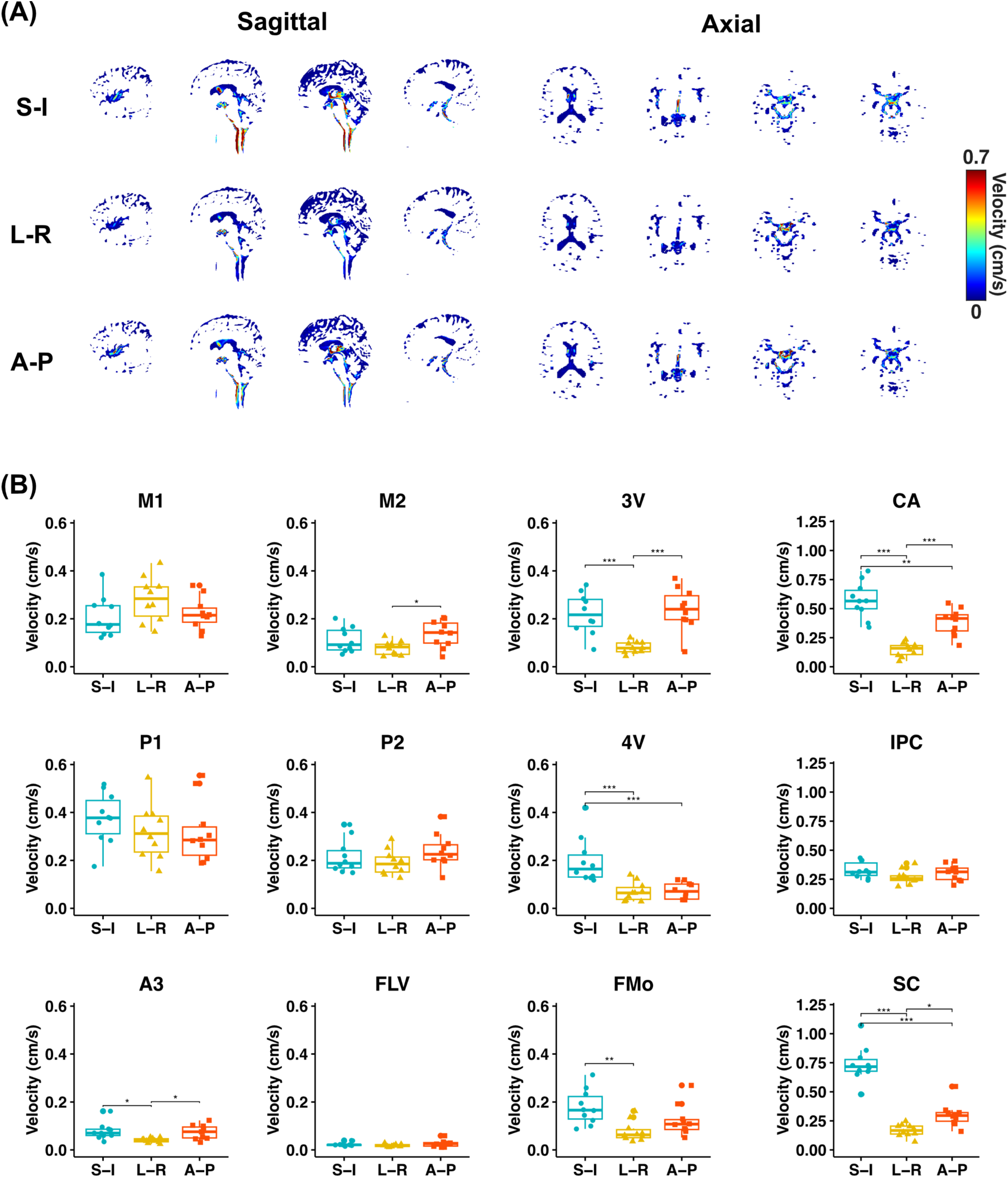
The VSSL-derived velocity maps for three different velocity-encoding directions. (A) The VSSL-derived velocity maps of S-I, A-P, and L-R directions for a representative healthy subject. (B) The boxplots of VSSL-derived velocity in each ROI across three orthogonal velocity-encoding directions. One-way ANOVA with post-hoc Tukey’s honestly significant difference tests for pairwise comparisons was used for comparison (* *p* < 0.05, ** *p* < 0.01, *** *p* < 0.001).

**Table 2.** Statistical results of VSSL-derived velocity along S-I, L-R, and A-P directions, and absolute velocity (N = 10).

| Velocity in ROIs (cm/s) |  | M1 | M2 | P1 | P2 | A3 | FLV | 3V | 4V | FMo | CA | IPC | SC |
| --- | --- | --- | --- | --- | --- | --- | --- | --- | --- | --- | --- | --- | --- |
| S-I | Mean | 0.208 | 0.111 | 0.376 | 0.215 | 0.078 | 0.024 | 0.221 | 0.198 | 0.179 | 0.572 | 0.329 | 0.735 |
|  | SD | 0.084 | 0.051 | 0.106 | 0.069 | 0.035 | 0.008 | 0.085 | 0.096 | 0.072 | 0.155 | 0.068 | 0.150 |
| L-R | Mean | 0.281 | 0.078 | 0.318 | 0.191 | 0.040 | 0.019 | 0.081 | 0.070 | 0.077 | 0.147 | 0.268 | 0.168 |
|  | SD | 0.093 | 0.029 | 0.112 | 0.052 | 0.010 | 0.005 | 0.024 | 0.039 | 0.042 | 0.059 | 0.061 | 0.053 |
| A-P | Mean | 0.222 | 0.136 | 0.315 | 0.240 | 0.074 | 0.026 | 0.240 | 0.073 | 0.123 | 0.386 | 0.306 | 0.302 |
|  | SD | 0.067 | 0.056 | 0.128 | 0.070 | 0.029 | 0.014 | 0.088 | 0.035 | 0.064 | 0.113 | 0.070 | 0.102 |
| Absolute Velocity | Mean | 0.416 | 0.194 | 0.589 | 0.378 | 0.116 | 0.041 | 0.338 | 0.224 | 0.232 | 0.711 | 0.525 | 0.819 |
|  | SD | 0.135 | 0.076 | 0.184 | 0.096 | 0.043 | 0.013 | 0.120 | 0.105 | 0.100 | 0.178 | 0.105 | 0.157 |

| *p*-value in ROIs |  | M1 | M2 | P1 | P2 | A3 | FLV | 3V | 4V | FMo | CA | IPC | SC |
| --- | --- | --- | --- | --- | --- | --- | --- | --- | --- | --- | --- | --- | --- |
| ANOVA |  | 0.123 | 0.033 | 0.432 | 0.251 | 0.007 | 0.268 | <0.001 | <0.001 | 0.003 | <0.001 | 0.140 | <0.001 |
| HSD Test | S-I vs L-R | n.s. | n.s. | n.s. | n.s. | 0.012 | n.s. | <0.001 | <0.001 | 0.002 | <0.001 | n.s. | <0.001 |
|  | S-I vs A-P | n.s. | n.s. | n.s. | n.s. | n.s. | n.s. | n.s. | <0.001 | n.s. | 0.003 | n.s. | <0.001 |
|  | L-R vs A-P | n.s. | 0.025 | n.s. | n.s. | 0.022 | n.s. | <0.001 | n.s. | n.s. | <0.001 | n.s. | 0.028 |

## Discussion

This study proposes the use of the VSSL MRI method to measure the CSF flow velocity within the ventricles and around the arteries in SAS. Modified from the sequence for measuring cerebral blood volume (32,49), the VSSL employed in this CSF study applied Fourier-transform-based inversion instead of Fourier-transform-based saturation pulse trains for higher CSF signals. Both the VSSL pulse train and the 3D-GRASE readout afford extended duration by taking advantage of the rather long CSF T_2_ (54). In contrast to previous velocity-selective arterial spin labeling studies which aimed to separate the signal of blood and static tissue into the pass band and stop band of the designed VSSL profiles, this work focuses on the transition band to linearly encode the flow velocity.

The CSF flow pattern observed in the ventricles is highly consistent with those observed using 3D amplified MRI (aMRI) (60) and other MRI methods (29,61). All methods demonstrated that the highest CSF flow occurs around the IPC, SC, and CA regions. The VSSL method can effectively measure CSF flow in the SAS along major arteries, including the MCA, ACA, and PCA. The average CSF flow velocity around the arteries in SAS is 0.339 ± 0.117 *cm*/*s*, which is close to and even higher than the flow velocity in the ventricles (0.309 ± 0.116 *cm*/*s*). However, the CSF flow along major veins, such as the superior sagittal sinus, is below the detection threshold of VSSL. Interestingly, there is no clear orientation dependence around most arteries in SAS, except for the A3 segment of ACA. This suggests that the CSF flow around the arteries in SAS is mainly turbulent. The flow velocity around the proximal arteries in SAS is higher than that of distal arteries, i.e., the velocity around M1 is higher than M2, and P1 is higher than P2 (Figure 7).

It is important to note that many diffusion-based MRI methods have been proposed to analyze CSF dynamics in the PVS (62), such as low b-value dynamic diffusion imaging (24), DTI-ALPS (23), and improved multi-directional diffusion-sensitized driven-equilibrium (iMDDSDE) (61). However, a major challenge in these experiments is suppressing the diffusion component, which can easily be mixed with flow measurements. It is evident in the iMDDSDE study, where signals can be seen in all components filled with CSF such as SAS. The diffusion component is comparable to the flow signal, as evidenced by the dynamic patterns of iMDDSDE over a cardiac cycle. The diffusion component can be separated from the CSF flow by utilizing the fact that flow strongly depends on cardiac movement, which is the principle behind the dynamic diffusion-weighted imaging (dDWI) method. (24) In the current study, we implemented a flow-compensated gradient pair to suppress the diffusion component. The b-value difference between the control and label in VSSL, with the parameters applied in this study, is relatively small ( Δ*b* = 2.21 *s* ∕ *mm*^2^), which is confirmed by the low VSSL signal in the cortex sulci (Fig. S2). In principle, the PVS in penetrating arteries, such as those within the basal ganglia and cortex, can be detected using VSSL with higher b-values and reduced Tseg, as the T_2_ in PVS may be significantly shorter than that of CSF in the ventricles. However, the b-value difference between the control and label would be too high to be neglected, requiring an improved b-value compensation approach to effectively suppress the diffusion component. Another key difference between iMDDSDE and VSSL is that the iMDDSDE signal follows a sinusoidal function relative to the flow velocity, making it challenging to distinguish between high and low flow velocities. In contrast, the VSSL signal is still proportional to the flow velocity beyond the V_cut_ as shown in Figure 6D. One more challenge with diffusion-based methods to study CSF flow is that changes in the apparent diffusion coefficient (ADC) can conflate flow changes with variations in perivascular fluid content, capillary perfusion, and even partial volume effect due to cerebral atrophy (63). The long-TE GRASE technique provides significant advantages by effectively suppressing signals from blood and tissue, which helps to mitigate these issues.

Although a few studies have been conducted specifically on the SAS (64–67), due to the difficulty in suppressing the diffusion component, most research has focused on the entire SAS. With efficient suppression of diffusion and 3D imaging, our study clearly delineates the entire SAS and reveals a pattern of the PVSAS, closely resembling the gadolinium-enhanced imaging results (16). However, whether high CSF flow is confined to the PVSAS or extends to other regions of the SAS requires further investigation. Our method offers a non-invasive tool to examine both the anatomy and CSF dynamics of the SAS system, and has the potential to delineate the newly discovered PVSAS system. Previous research (16) has reported the PVSAS as a donut-shaped form around the arteries, although the exact diameter of the PVSAS was not specified. Based on prior studies of the PVS, normal PVS is generally smaller than 2 mm in size (68). Another study showed that the PVS appears linear when imaged parallel to the course of the vessel, with a diameter generally smaller than 3 mm when imaged perpendicular to the vessel’s course (44). Additionally, the VSSL signal around the major arteries in the SAS exceeds 2mm, as demonstrated in Supplementary Figure S3. Thus, we confined our study to the area around the arteries within 1 mm, which aims to mainly focus on the PVSAS system.

PC-MRI exhibits a relatively high noise level, with a mean velocity of 0.45 cm/s measured in the parenchyma. Due to its high baseline noise, PC-MRI faces challenges in accurately measuring low flow rates and is predominantly suitable for regions with high velocities, such as the IPC, PPC, and SC. This limitation significantly restricts PC-MRI’s utility in measuring velocities around most arteries in SAS and PVS. Hence, our calibration relies on regions with high velocities, including the IPC, PPC, and SC, assuming a linear correlation between PC-MRI velocity and normalized VSSL signal. Notice that the linear assumption may be only valid for complex flow patterns and may not apply to simpler, single-layer flow. Given the predominance of complex flow in physiological settings, this assumption is valid and is further supported by the PC-MRI calibration study (Figure 6D). With a laminar flow model, the normalized VSSL signal is a linear function of the flow velocity when it is less than 0.8 cm/s.

In this study, we measured the average CSF flow velocity throughout the entire cardiac cycle. Due to the relatively long acquisition times required for the VSSL module and 3D-GRASE readout, it is challenging to measure the velocity waveform within a single cardiac period, as typically done with PC-MRI. While it is still possible to obtain the velocity waveform using a retrospective gating approach similar to that used in the iMDDSDE (61) and dDWI (24) methods, this would significantly increase the total duration of the experiment.

## Conclusions

This study successfully employed the VSSL MRI method to measure the CSF flow rate around the arteries in SAS and ventricles. The observed CSF flow patterns in the ventricles were highly consistent with those obtained using 3D amplified MRI (aMRI) and other advanced MRI techniques, verifying the reliability of the VSSL method. The VSSL method demonstrated its capability in measuring CSF flow along major arteries such as the MCA, ACA, and PCA in SAS. Interestingly, the CSF movement along major arteries in SAS showed weak orientation dependence, suggesting a complex driving force for CSF flow. This study underscores the potential of VSSL MRI as a non-invasive tool for investigating CSF dynamics, offering significant insights into the understanding of CSF circulation in both healthy and pathological conditions. Future studies could benefit from the application of improved diffusion compensation approaches to further suppress diffusion components and enhance the accuracy of CSF flow detection in smaller arteries.

## Data Availability

The scripts and code used in this study will be shared publicly upon publication of the article. The brain imaging data used in this study are available on request according to JHU material transfer agreement (MTA).

## Acknowledgments

This work was supported by P41EB031771, R01HL149742, R01AG080104 and R21AG074978. The authors thank D, Mr. Joseph S. Gillen, Mrs. Terri Lee Brawner, Ms. Kathleen A. Kahl, and Ms. Ivana Kusevic for experimental assistance.

---

Figure S1. Simulated Mz-velocity responses for the VSSL pulse train with segment times (T_seg_) as 10 (A), 20 (B), 30 (C), 40 (D), and 50 (E) ms. Velocity-sensitive and velocity-compensated profiles are represented by solid and dashed blue lines, respectively. The cut-off velocity (V_cut_) is delineated at the first intersection where ΔM = 1, highlighted by vertical black dashed lines. Parameters used in the simulation, such as number of segments (8) and G_max_ (40 mT/m), are consistent with those employed in our study.

Figure S2. The VSSL images (top row) and M0 images (bottom row) with different TEs (TE = 816 and 39 ms).

Figure S3. The cross-sections of the M1 (A), M2 (B), P1 (C), and P2 (D) arteries. The blood vessels are shown in red, and the VSSL signal is displayed in green. Insert images show the magnified views of the PVSAS around arteries.

