## Supplementary Figure for "Cerebrospinal Fluid Flow within Ventricles and Subarachnoid Space Evaluated by Velocity Selective Spin Labeling MRI"

### Supporting Information:

#### Simulated velocity responses for the VSSL pulse train with different segment times

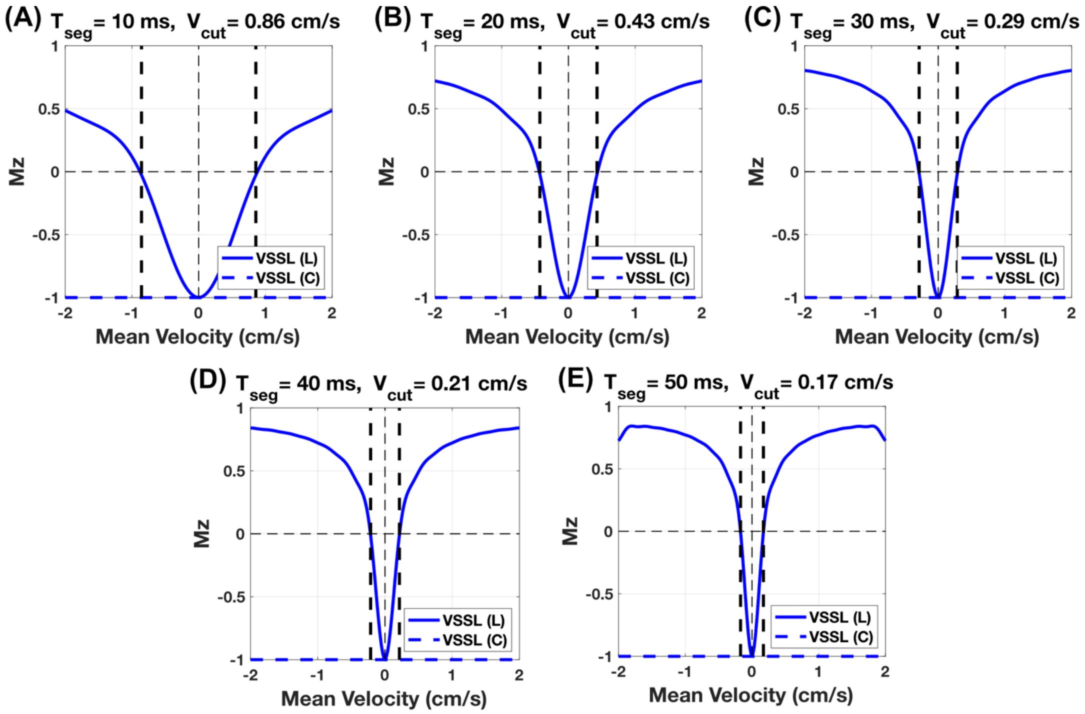


Figure S1. Simulated Mz-velocity responses for the VSSL pulse train with segment times ($T_{seg}$) as 10 (A), 20 (B), 30 (C), 40 (D), and 50 (E) ms. Velocity-sensitive and velocity-compensated profiles are represented by solid and dashed blue lines, respectively. The cut-off velocity ($V_{cut}$) is delineated at the first intersection where $\Delta M = 1$, highlighted by vertical black dashed lines. Parameters used in the simulation, such as number of segments (8) and $G_{max}$ (40 mT/m), are consistent with those employed in our study.

**Comparison of the VSSSL and M0 images acquired with different TE values**

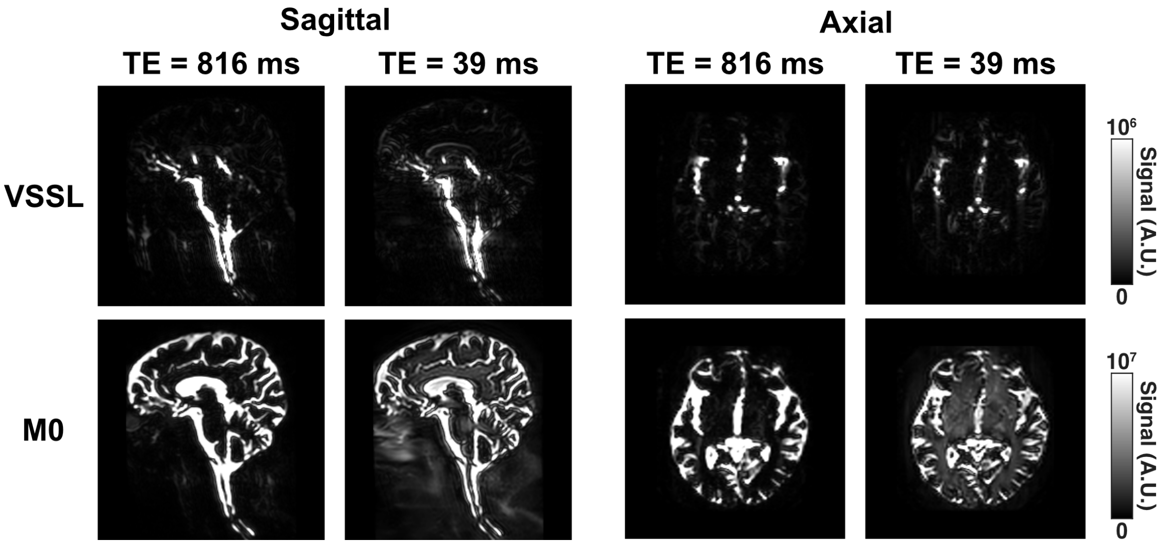


Figure S2. The VSSL images (top row) and M0 images (bottom row) with different TEs (TE = 816 and 39 ms).

**The cross-sections of the major arteries acquired with VSSL and MRA**

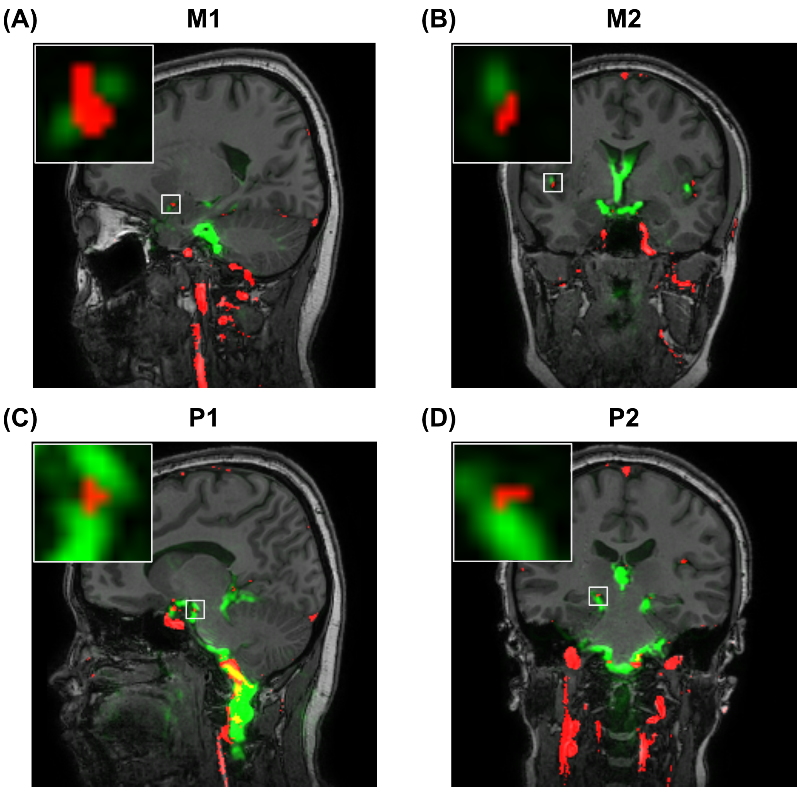

Figure S3. The cross-sections of the M1 (A), M2 (B), P1 (C), and P2 (D) arteries. The blood vessels are shown in red, and the VSSL signal is displayed in green. Insert images show the magnified views of the PVSAS around arteries.
